# A novel demographic-based model shows that intensive testing and social distancing are concurrently required to extinguish COVID-19 progression in densely populated urban areas

**DOI:** 10.1101/2020.06.23.20138743

**Authors:** Mario Moisés Alvarez, Grissel Trujillo-de Santiago

## Abstract

We present a simple epidemiological model that includes demographic density, social distancing, and efficacy of massive testing and quarantine as the main parameters to model the progression of COVID-19 pandemics in densely populated urban areas (i.e., above 5,000 inhabitants km^2^). Our model demonstrates that effective containment of pandemic progression in densely populated cities is achieved only by combining social distancing and widespread testing for quarantining of infected subjects. Our results suggest that extreme social distancing without intensive testing is ineffective in extinguishing COVID-19. This finding has profound epidemiological significance and sheds light on the controversy regarding the relative effectiveness of widespread testing and social distancing. Our simple epidemiological simulator is also useful for assessing the efficacy of governmental/societal responses to an outbreak.

This study also has relevant implications for the concept of smart cities, as densely populated areas are hotspots that are highly vulnerable to epidemic crises.

## Introduction

COVID-19 has clearly illustrated that we were unprepared for the second pandemic of the 21^st^ century. By the third week of July 2020, the official cumulative number of infected persons worldwide was more than 14,000,000, with a death toll higher than 600,000. COVID-19 has not recognized frontiers; it started in China, migrated to Iran, and formed a later epicenter in Italy and Spain. It then extended to France, England, Germany, and other European Countries. Now, COVID-19 has a strong presence in Las Americas, the current epicenter of the pandemics, mainly in the USA, Brazil, and Mexico[1,2]. India just became the third country with the most officially confirmed cases (after USA and Brazil) currently reports more than 1,000,000 cases of COVID-19. Precisely India, USA, Brazil and México concentrate many of the most densely populated cities in the planet (i.e., New York, Kalcota, Bangalore, Sao Paulo, Rio de Janeiro, and México City). Among all affected territories, pandemic COVID-19 has encountered containment responses ranging from aggressive to weak. Some of these responses, namely the ones that appear to be more successful, have been based on a combination of social distancing and massive testing and selective quarantine[3]. Social distancing effectively decreases the progression of the disease essentially by decreasing the demographic density, while massive testing enables the identification of infected subjects and opportune quarantining[4] (which is technically a version of selective social distancing). However, discussion persists regarding the relative effectiveness of countermeasures such as social distancing or massive testing, and countries have used social distancing and testing effort with widely different emphasis to face COVID-19. Countries like South Korea have based their strategy of containment on strict social distancing and massive testing in their open populations[5]. Spain initially based its strategy on gradual social distancing. Later, it significantly increased its diagnostic efforts to plateau the pandemic. England started by using testing mainly for epidemiological recording, but later significantly scaled up its efforts to aggressively diagnose its population. By contrast, Mexican officials have openly declared that massive testing and quarantine have no significant value as a countermeasure for COVID-19 progression and that diagnostics is mostly informative; Mexico has been one of the countries with the fewest diagnostic tests conducted per 100,000 inhabitants[6].

The benefit of social distancing and massive diagnostics has not only been a subject of controversy[7], but assessing the potential benefit has also been challenging.

Here, we introduce a simple mathematical model with a formulation that explicitly considers demographic variables (i.e., the demographic density and the total population) to calculate the progression of COVID-19 in urban areas. The model also considers the effectiveness of social distancing measures and of massive testing for expeditious identification and quarantining of infective subjects as inputs. We present a wide range of simulation scenarios for “representative” urban areas and show that both social distancing and widespread and effective testing should be combined for effective containment of the pandemic advance in densely populated urban areas.

### Model formulation

We developed a very simple epidemiological model for the propagation of COVID-19 in urban areas (Figure 1A; S1 File). The model considers two variable populations of individuals, infected (X) and retrieved (R). The cumulative number of infected patients (X) is the total number of subjects among the population that have been infected by SARS-CoV-2. The number of retrieved patients should be interpreted as the number of individuals that have been retrieved from the general population and are not contributing to the propagation of COVID-19. Retrieved subjects include subjects who have recovered from the infection and do not shed virus, quarantined individuals, and deceased patients. Importantly, the model assumes that infection results in (at least) short-term immunity upon recovery. This assumption is based in experimental evidence that suggests that rhesus macaques that recovered from SARS-CoV-2 infection could not be reinfected[8]. However, the acquisition of full immunity to reinfection has not been confirmed in humans, although it is well documented for other coronavirus infections, such as SARS and MERS[9,10].

**Figure 1.**
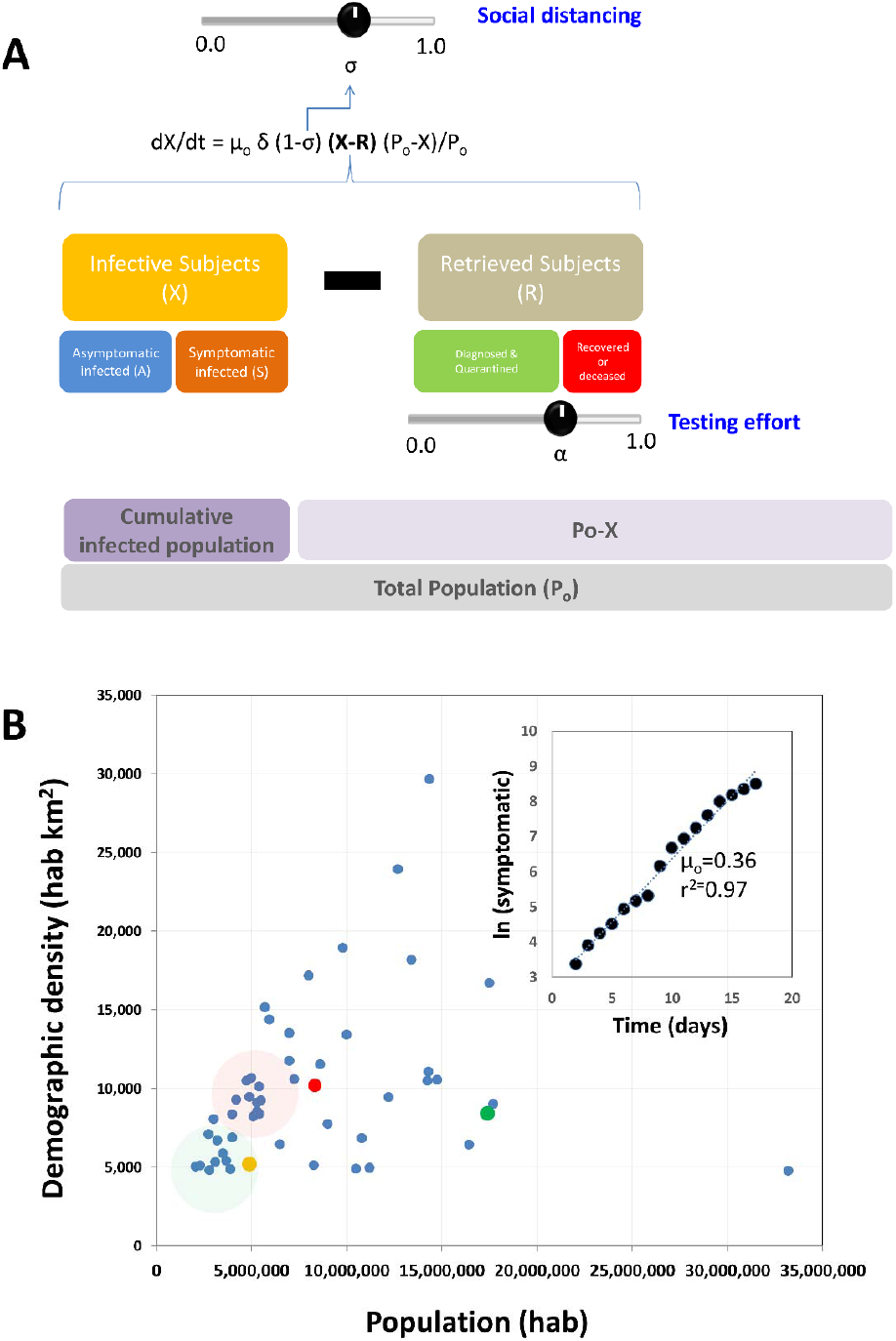
Model formulation and scenarios. (A) Schematic representation of the model. (B) Population and demographic density of the world largest cities.Two clusters of cities, referred here as Type I (green cluster) and Type II (red cluster) are indicated in a plot of population versus demographic density. The 51 largest cities in the world, according to www.citymayors.com/, are included[14]. Madrid Spain is indicated in yellow (•), New York City in red (•), and Mexico City in green (•). (b) Natural log of the number of symptomatic versus time in Madrid from March 2 to March 17 (initial stage of the pandemics in Spain). The initial rate of infection (µ_o_) can be calculated from the slope of this straight line.

Two set of parameters, demographic and clinical/epidemiological, determine the interplay between these two main populations and other subpopulations that include asymptomatic infected (A), symptomatic infected (S), and deceased (D). Clinical parameters include an intrinsic infection rate constant (µ_o_) that is calculated from the initial stage of the pandemic in that particular region, the fraction of asymptomatic patients (a), the delay between the period of viral shedding by an infected patient (delay_r), the period from the onset of shedding to the result of first diagnosis and quarantine in the fraction of patients effectively diagnosed (delay_q), and the fraction of infected patients effectively diagnosed and retrieved from the population (α). Demographic parameters include the population of the region (P_o_), the relative demographic density (δ), the social distancing (σ), and the fraction of infected individuals retrieved from the population due to massive and effective testing (α) (Figure 1A). The model is based on a set of two simple differential equations.

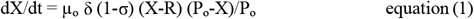

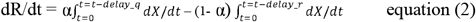

The first equation of the set (equation 1) states that the rate of accumulation of infected habitants (symptomatic and asymptomatic) in an urban area (assumed to be a closed system) is proportional to the number of infective subjects (X-R) present in that population at a given point and the fraction of the population susceptible to infection ((P_o_-X)/P_o_). Note that the number of infective subjects is given by the difference between the accumulated number of infected subjects (X) and the number of retrieved subjects (R). The fraction of the susceptible population decreases over time as more inhabitants in the community get infected. The proportionality constant in equation 1 (µ_o_) is an intrinsic rate of infection that is weighted by the population density (δ) in that urban area, and the effective fractional reduction of social distancing on the population density (1- σ).

The second equation (equation 2) describes the rate at which infected patients are retrieved from the infective population. Eventually, all infected subjects are retrieved from the population of infected individuals, but this occurs at distinctive rates. A fraction of infected individuals (α) is effectively retrieved from the general population soon after the onset of symptoms or after a positive diagnosis. Another fraction of infected subjects (1- α) is not effectively retrieved from the population until they have recovered or died from the disease. Therefore, in our formulation, the overall rate of retrieval (dR/dt) has two distinctive contributions, each one associated with different terms on the right-hand side of equation 2. The first term accounts for the active rate of retrieving infected patients through the diagnosis and quarantine of SARS-CoV-2 positive subjects. For this term, the delay from the onset of virus shedding to positive diagnosis and quarantine (delay_q) is considered short (*i*.*e*., between two or three days), to account for a reasonable time between the positive diagnosis and the action of quarantine. In our model formulation, this term is represented by (1-α). A second term relates to the recovery or death of infected patients (symptomatic or asymptomatic) and is represented by the integral of all infected subjects recovered or deceased from the onset of the epidemic episode in the region, considering a delay of 21 days (delay_r), which accounts for the average time of recovery of an infected individual. Note that the simultaneous solution of equations 1 and 2 is sufficient to describe the evolution of the number of asymptomatic individuals (A), symptomatic individuals (S), and deceased patients (D) through the specification of several constants and simple relations.

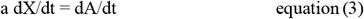

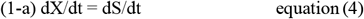

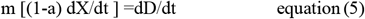

Here, **a** is the fraction of asymptomatic subjects among the infected population, (1-**a**) is the fraction of infected individuals that exhibit symptoms, and **m** is the mortality rate expressed as a fraction of symptomatic individuals.

### Selection of relevant epidemiological parameters

As with any epidemiological model, this model relies on some basic assumptions that must be sustained in clinical or epidemiological data (Table 1). We now briefly discuss the assumptions that were made and the rationale behind the relevant values of the parameters of the model: the fraction of asymptomatic infected, the average time to recover, the fraction of symptomatic patients that would require hospitalization, and the average time of bed occupancy per hospitalized patient.

**Table 1.** Epidemiological data and parameter values used in the model.

| Parameter | Definition (units) | Value or [range] | Reference/source |
| --- | --- | --- | --- |
| $\mu_o$ | Intrinsic rate of infectivity of COVID-19 ($\text{day}^{-1}$)<br>Calculated from confirmed COVID-19 cases during the first stage of the pandemic at Madrid, Spain (Ministry of Health, Spain):<br>(https://www.mscbs.gob.es) | 0.36 | Calculated |
| $\delta$ | Relative demographic density<br>(normalized respect to the demographic density of Madrid, Spain) (dimensionless)<br>Parameter provided by model user to simulate a scenario | [0.75-2.00] | User defined |
| $\sigma$ | Social distancing (dimensionless)<br>Parameter provided by model user to simulate a scenario | [0-1] | User defined |
| (1-σ) | Fractional reduction in activity in the city due to social distancing [range:0-1] | [0-1] | Calculated from $\sigma$ |
| $P_o$ | Total population of the city (hab)<br>Provided by model user from actual census data; (i.e.; www.citymayors.com) | [3,500,000-] | [14] |
| delay_q | Average number of days between infection and positive diagnostics and quarantine (days) | 5 | assumed |
| delay_r | Average number of days between infection and effective recovery or dead (days) | 21 | [15], [16]. |
| a | Fraction of asymptomatic subjects among infected.<br>Inferred from serological study conducted in NYC: (www.cnn.com) | 0.90 | [11] |
| (1-a) | Fraction of symptomatic subjects among infected | 0.10 | [11] |
| m | Mortality rate expressed as a fraction of symptomatic individuals.<br>Continuously updated by Johns Hopkins University (Coronavirus Research Center).<br>(https://coronavirus.jhu.edu/) | 0.069 | (2) |

The fraction of asymptomatic infected is one of the critical inputs to the model; it determines the final and maximum feasible threshold of symptomatic infected. However, the current evidence is not yet sufficient to support a conclusive value for this parameter. Nevertheless, a recent serological study conducted in New York City (NYC) found anti-SARS-CoV-2 IGGs among 21.2% of the population[11] (www.cnn.com), and this result is consistent with previous information related to massive epidemic episodes. For instance, in the context of the pandemic influenza A/H1N1/2009, up to 20–40% of the population in urban areas (i.e., Monterrey, México, and Pittsburgh, USA) [12,13] exhibited specific antibodies regardless of experiencing symptoms, while the fraction of confirmed symptomatic infections was lower than less than 10%. This serological result based exclusively on information from NYC suggests that more than 90% of exposed New Yorkers (∼91.4%) were asymptomatic or exhibited minor symptoms. Based on this (still unpublished) data, we assumed a symptomatic fraction of only 10% in the calculations and forecasts presented here.

In addition, the average time of sickness was set at 21 days in our simulations, as this is within the reported range of 14 to 32 days[15], with a median time to recovery of 21 days[16]. Viral shedding can last for three to four weeks after the onset of symptoms, with a peak at day 10-11.[17] Therefore, we assume that all those infected not quarantined could continue to transmit the virus until full recovery (21 days). Similarly, asymptomatic patients are only removed from the pool of susceptible persons after full virus clearance. Note that, in the current version of our model, asymptomatic patients are considered part of the population capable of transmitting COVID-19; reported evidence that suggests that asymptomatic subjects (or minimally symptomatic patients) may exhibit similar viral loads[18] to those of symptomatic patients and may be active transmitters of the disease[19,20].

The average fraction of deceased patients worldwide is estimated as 0.064 of those infected 21 days before. This mortality percentage (case fatality rate) lies within the range reported in the recent COVID-19 literature[21–24]. The time lapse of 14 days between the onset of disease and death was statistically estimated by Linton et al. in a recent report [25]. We also consider that the average time for bed occupancy of hospitalized patients is 14 days. The estimated average hospitalization stays range from 9.3 to 13 days in the United States [26] and China[27,28], but much longer stays have been reported in intensive care units in Italy (20 to 25 days)[29]. Anecdotal data collected in México suggests that hospitalization stays of at least two weeks are a more accurate figure for Latin American societies.

### Definition of representative scenarios

We aimed to reproduce representative settings for COVID-19 progression; therefore, we selected two hypothetical but realistic urban scenarios. Figure 1B shows the most densely populated cities in the world (according to information concentrated in www.citymayors.com)[14] represented by dots in a bidimensional plot of demographic density versus population. Two clusters of cities can be readily identified. The first group of 5 to7 important cities in the world (i.e, Porto Alegre, Ankara, Athens, Guadalajara, Monterrey, Barcelona) have populations between 3.0 and 3.7 million inhabitants and a demographic density of ∼5,000 inhabitants per square kilometer (hab km^2^). A second group is centered on the coordinates of a population of 10 million citizens and a demographic density of 10,000 hab km^2^ (i.e., Baghdad, Ho Chi Minh, Bangalore, Tianjin, Kinshasa, Hyderabad). Based on this analysis, we centered our estimates of COVID-19 progression in these two classes of “representative” cities (3.5 million inhabitants, 5,000 hab km^2^; and 10 million citizens, 10,000 hab km^2^).

We also based our estimates of µ_o_, the intrinsic rate of COVID_19 propagation, on data extracted from the local dynamics of the pandemics in Madrid (a city in our first category) and New York City (a city in our second category). These two cities also belong to a limited number of cities that have generated reliable datasets on the local progression of the number of COVID-19 positive cases over time. We calculated the value of µ_o_ (*i*.*e*., the intrinsic rate of infectivity of SARS-CoV2 before interventions) by assuming that the initial rate of propagation is d(X)/dt = µ_o_ [X], where [X] is the number of initially infected subjects. We then simply calculate the intrinsic rate of infection from the initial slope of a plot of ln [X] *vs* time, which is a usual procedure for calculation of intrinsic growth rates in cell culture scenarios under the assumption of first order rate growth dependence. Following this rationale, we set µ_o_ for all our simulations. Consistently, we noted that the initial rate of propagation observed in NYC, a city with twice the demographic density of Madrid, doubles this µ_o_ value. Therefore, we take the demographic density of Madrid as a reference (5,185 hab km^2^) and assume that multiplying µ_o_ by the normalized demographic density (with respect to that in Madrid) is a reasonable procedure for adapting the model to any urban area.

### Effect of social distancing and massive testing

Social distancing has been regarded as the one of the most effective buffering measures for containment of local COVID-19 epidemics[30–33]. However, the effectiveness of massive testing, either alone or in combination with social distancing, has been evaluated formally in a limited number of contributions[34–36].

First, we conducted simulations in which we evaluated the independent impact of different degrees of social distancing and massive testing in a Type I city (3.5 million inhabitants and medium demographic density). Figure 2A shows the impact of different degrees of social distancing at a fixed and basal value of massive testing. In this simulation, a basal value of social distancing (α=0.1) means that only 10% of the infected patients are diagnosed and quarantined, while the rest of the infected subjects continue active until recovery. This strategy is consistent with that adopted by countries that diagnosed essentially only those subjects who were symptomatic and asked for medical assistance (i.e., México, Chile, and Bolivia, with fewer than 2 tests per confirmed case [6]). The pandemic progression (number of cumulative symptomatic cases, new infections per day, and bed occupancy per day) is indicated with grey curves for a reference scenario with no social distancing and basal level of testing. Higher degrees of enforcement of social distancing (i.e., such that the demographic density is effectively reduced by 20, 40, and 60%) are presented with blue, green, and red lines, respectively. Levels of social distancing of 20% and 40% delay the pandemic curve by 15 and 30 days, whereas the pandemic progression is successfully buffered only when social distancing effectively reduces demographic density (and therefore activity) by 60% for extended times (i.e., for 6 months). Although eventually effective, this strategy of drastic and prolonged social distancing may be unsustainable or drastically damaging for the economy of low and medium income societies[32].

**Figure 2.**
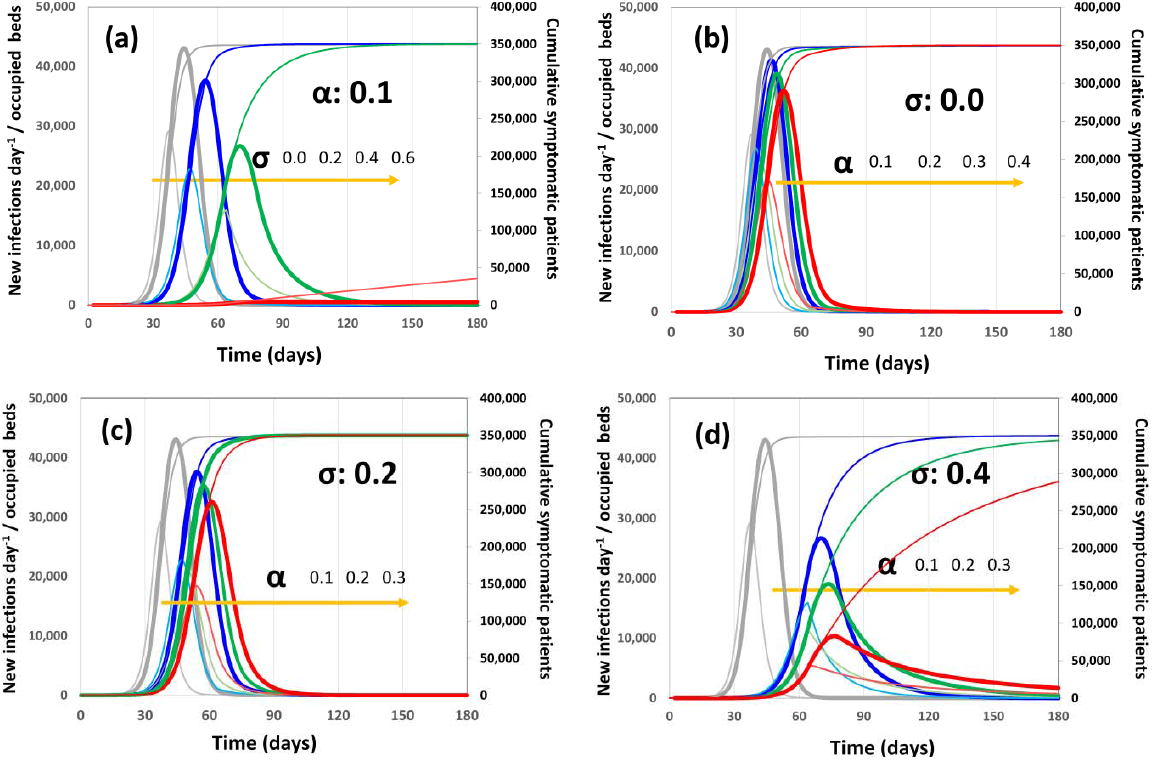
Pandemic progression scenarios for a Type I city (3.5 × 10^6^ citizens) with a medium demographic density (5,500 inhabitants km^-2^). The number of new infections per day (thin and light line curves), the bed occupancy (thick line curves) and the cumulative number of infections (thin cumulative curve) are presented for each scenario. (a) Scenario I: basal level of testing (α=0.1) and increasingly higher levels of social distancing: σ=0.0 (gray), σ=0.2 (blue), σ=0.4 (green), and σ=0.6 (red). (b) Scenario II: social distance not enforced (σ=0.0) and increasing values of testing α=0.1 (gray), α=0.2 (blue), α=0.3 (green), and α=0.4 (red). (c) Scenario III: social distance modestly enforced (σ=0.2), and increasing values of testing, α=0.1 (blue), α=0.2 (green), and α=0.3 (red). (d) Scenario IV: social distance moderately enforced (σ=0.4) and increasing values of testing α=0.2 (blue), α=0.3 (green), and α=0.4 (red). Gray curves correspond to the pandemic progression with minimum intervention (no social distancing and α=0.1).

Figure 2B presents predictions for scenarios where no social distancing is implemented, but where the level of diagnostic testing is increased from a basal value (10% of infected are diagnosed and quarantined) to situations were 20, 30, and 40% of infected patients (symptomatic and asymptomatic) are diagnosed within the first 3 days of viral shedding and quarantined. These levels of massive testing, in the absence of social distancing enforcement, are not sufficiently effective to buffer the pandemic.

Figure 2C shows a scenario in which social distancing is set at 20% and different testing emphasis is applied. As before, trends related to the reference scenario of low testing and no social distance enforcement are indicated in grey. The combination of moderate social distancing and testing renders better results than any one of the two strategies independently applied. When social distancing is elevated to a 40% and combined with more intensive testing efforts, the epidemic peak is dramatically delayed. A value of 40% social distancing in combination with an effort to identify and quarantine 30% of newly infected subjects delays the peak of maximum bed occupancy from day 25 to day 75 and lowers the highest demand of beds from 75,000 to fewer than 30,000. Increased testing at 40% social distancing further contributes to extinguish the epidemic peak.

We also estimated the pandemic progression under the same set of scenarios for a densely populated city (Type II: 5×10^6^ citizens and 10,000 hab km^2^). The results similarly indicate that only a combined strategy of social distancing and scaled-up testing and quarantine may effectively control the pandemic progression due to “selective social distancing”. However, the higher population leads to increases in the number of cases in the same time frame (let us say, the first 120 days from pandemic onset) and in the maximum threshold of symptomatic cases. The higher demographic density also causes a higher rate of transition. Therefore, the containment strategies need to be stronger than those required for Type I cities. For example, in the absence of intensified testing, the degree of social distancing required to buffer the pandemic is much higher in a Type II city (α=0.75) than in a type I city (α=0.60). Similarly, only aggressive social distancing interventions combined with intensified testing, *i*.*e*., (σ=0.6, α=0.3) or (σ=0.7, α=0.5) can mitigate the pandemic progression in larger or denser cities (Type II). A summary of the results for different combinations of scenarios is presented in Tables 2 and 3.

**Table 2.** Effect of social distancing and testing in a city with a demographic density of 5,500 inhabitants/km^2^ and a population of 3.5 × 10^6^ persons).

| | | $\alpha$ | | | | | | | | |
| --- | --- | --- | --- | --- | --- | --- | --- | --- | --- | --- |
|  |  | 0.10 | 0.20 | 0.30 | 0.40 | 0.50 | 0.60 | 0.70 | 0.80 | 0.90 |
| 0.00 | peak @ day: | 38.00 | 39.00 | 42.00 | 45.00 | 50.00 | 56.00 | 64.00 | 70.00 | 100.00 |
|  | infected after 120 days: | 349037.52 | 349072.31 | 349410.58 | 349731.45 | 349801.94 | 349008.93 | 323718.47 | 134600.03 | 5963.75 |
|  | peak of infection (hab/day) | 29155.11 | 26837.36 | 24343.10 | 21633.49 | 18652.11 | 15311.81 | 11469.93 | 2849.59 | 90.84 |
|  | maximum bed occupancy: | 43076.11 | 41297.81 | 39084.22 | 36297.24 | 32740.85 | 28128.49 | 20667.05 | 5774.46 | 190.73 |
| 0.25 | peak @ day: | 51.00 | 54.00 | 59.00 | 64.00 | 65.00 | 70.00 | 81.00 | 90.00 | 90.00 |
|  | infected after 120 days: | 349994.09 | 349980.90 | 349723.77 | 342776.43 | 274280.90 | 122542.87 | 24923.94 | 2614.45 | 248.17 |
|  | peak of infection (hab/day) | 21114.08 | 19067.32 | 16862.56 | 14462.14 | 8280.49 | 2397.63 | 398.56 | 37.26 | 2.59 |
|  | maximum bed occupancy: | 35889.86 | 33437.60 | 30474.06 | 25109.72 | 14935.49 | 4900.12 | 833.91 | 78.27 | 5.45 |
| 0.5 | peak @ day: | 67.00 | 67.00 | 76.00 | 81.00 | 82.00 | 86.00 | 88.00 |  |  |
|  | infected after 120 days: | 175936.83 | 91893.77 | 38845.99 | 13861.23 | 4332.07 | 1227.77 | 341.99 |  |  |
|  | peak of infection (hab/day) | 3803.14 | 1649.97 | 632.51 | 215.77 | 64.49 | 17.03 | 4.18 |  |  |
|  | maximum bed occupancy: | 7523.66 | 3411.07 | 1325.73 | 452.39 | 135.43 | 35.79 | 8.78 |  |  |
| 0.6 | peak @ day: | 77.00 | 81.00 | 83.00 | 86.00 |  |  |  |  |  |
|  | infected after 120 days: | 18919.36 | 8432.49 | 3534.87 | 1408.84 |  |  |  |  |  |
|  | peak of infection (hab/day) | 298.44 | 129.46 | 52.56 | 20.02 |  |  |  |  |  |
|  | maximum bed occupancy: | 625.81 | 271.66 | 110.40 | 42.07 |  |  |  |  |  |
| 0.7 | peak @ day: | 88.00 |  |  |  |  |  |  |  |  |
|  | infected after 120 days: | 1147.06 |  |  |  |  |  |  |  |  |
|  | peak of infection (hab/day) | 16.38 |  |  |  |  |  |  |  |  |
|  | maximum bed occupancy: | 34.42 |  |  |  |  |  |  |  |  |

(\*) Conditions at which the progression can be controlled or extinguished are indicated in light and intense green, respectively.

**Table 3.**
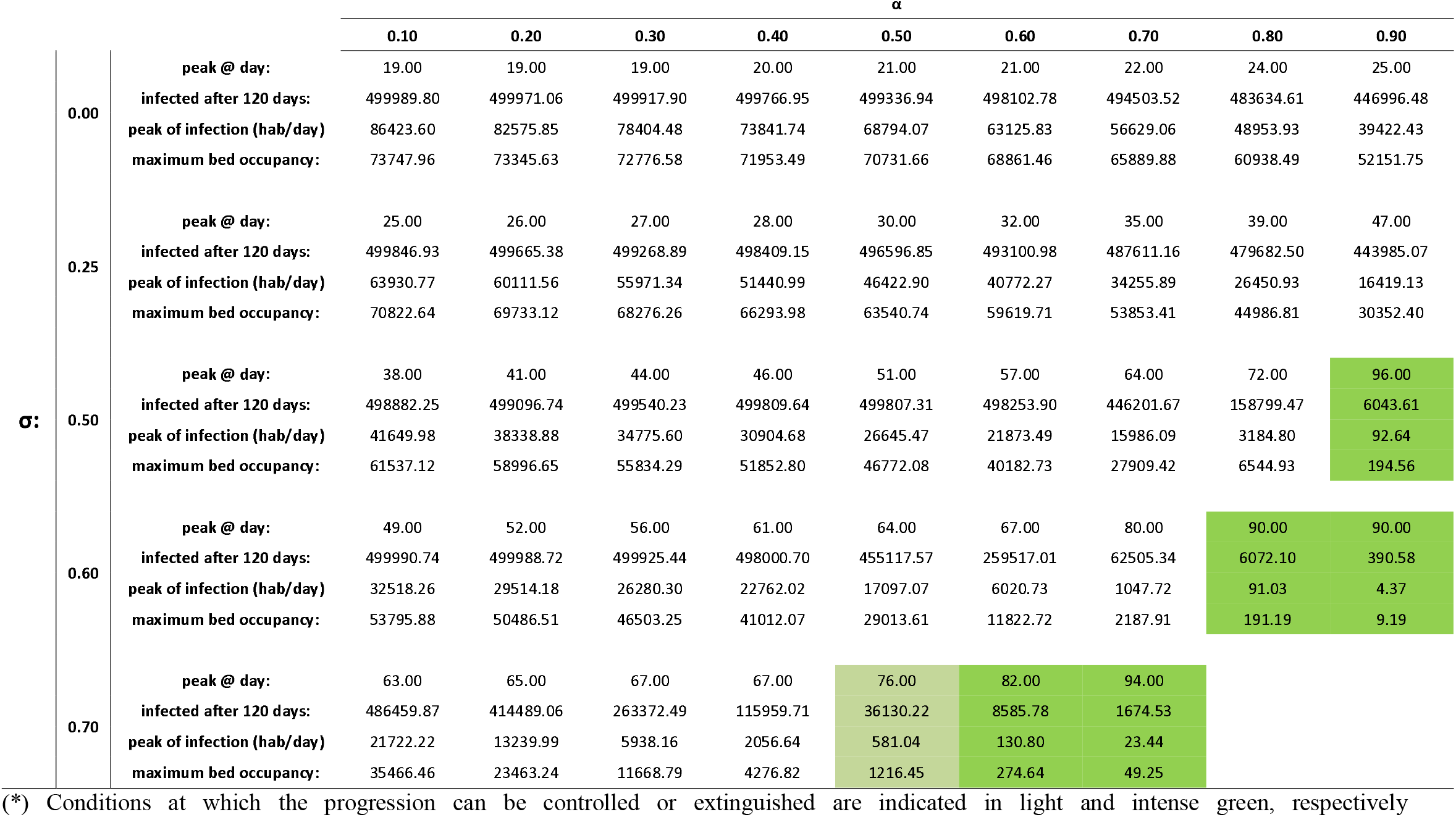
Effect of social distancing and testing in a city with a demographic density of 10,000 inhabitants/km^2^ and a population of 5.0 × 10^6^ persons).

We present scenarios for both moderately and densely populated cities (Type I and II). Four different indicators are calculated for each scenario, including the day of the epidemic peak, the number of new infection cases at the epidemic peak, the cumulative number of symptomatic infections after 120 days of the local pandemic onset (4 months), and the maximum bed occupancy.

**Figure 3.**
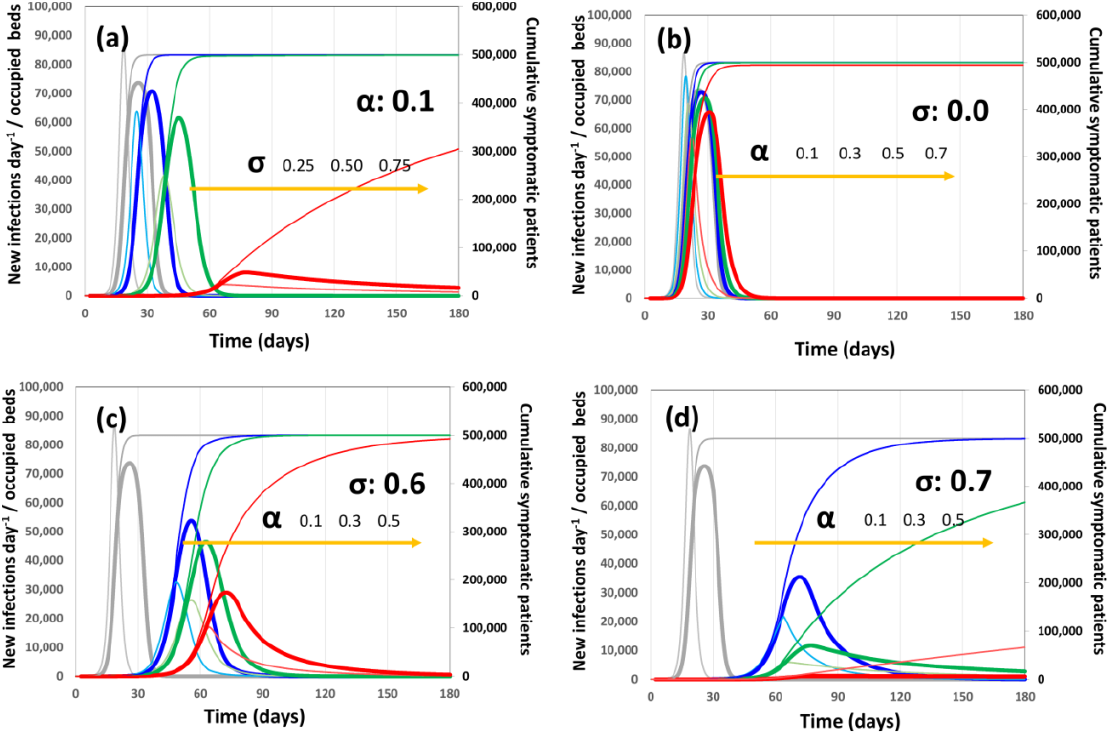
Pandemic progression scenarios for a Type II city (5.0 × 10^6^ citizens) with a high demographic density (10,000 inhabitants km^-2^). The number of new infections per day (thin and ligth line curves), the bed occupancy (thick line curves) and the cumulative number of infections (thin cumulative curve) are presented for each scenario. (a) Scenario I: basal level of testing (α=0.1) and increasingly higher levels of social distancing: σ=0.00 (gray), σ=0.25 (blue), σ=0.50 (green), and σ=0.75 (red). (b) Scenario II: social distance not enforced (σ=0.0) and increasing values of testing α=0.1 (gray), α=0.3 (blue), α=0.5 (green), and α=0.7 (red). (c) Scenario III: social distance enforced (σ=0.6) and increasing values of testing: α=0.1 (blue), α=0.3 (green), and α=0.5 (red). (d) Scenario IV: social distance moderately enforced (σ=0.7) and increasing values of testing: α=0.1 (blue), α=0.3 (green), and α=0.5 (red). Gray curves correspond to the pandemic progression with minimum intervention (no social distancing and α=0.1).

Next, we investigated the fidelity of the model prediction by simulating the advance of the pandemic in Madrid, NYC, and Mexico City. The number of new infections per day in these three cities has been made available by government officials [37–39]. Interestingly, the demographic characteristics of these major cities exhibit important differences, as well as the types of counter-measures adopted to contain COVID-19.

Madrid (a Type-I city) and NYC (nearly a Type-II city) have practically extinguished the pandemic. Therefore, the position and magnitude of the pandemic peak offers valuable information to estimate values of the magnitude of social distancing (σ) and diagnostic effort (α). In contrast, Mexico City has not reached the pandemic peak yet, and struggles to contain COVID-19 in a challenging demographic situation. We found sets of parameters that properly describe the evolution of COVID-19 in each of these cities, despite the obvious differences in the behavior of each progression curve (Figure 4). For Madrid, our simulations suggest that social distancing measures have achieved a degree of 58% of reduction in population density (α=0.58). However, even considering these relatively higher values of social distancing, an aggressive testing program (α=0.50) was needed to bend the curve at a cumulative number of ∼70,000 cases and reduce the emergence of new cases to the current levels (less than 10 per day). This implies that overall approximately 50% of the active infected subjects were found and quarantined through testing.

**Figure 4.**
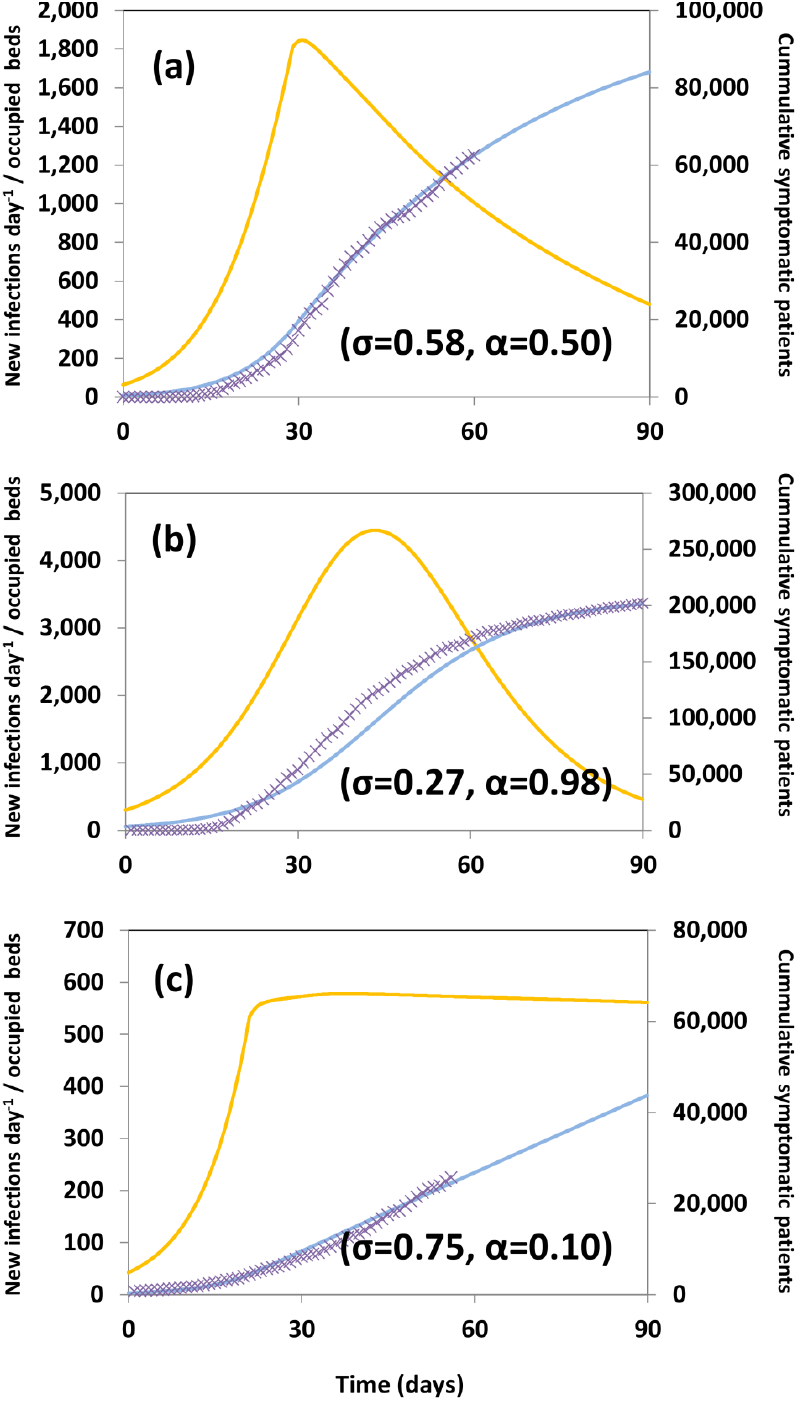
Progression of the COVID-19 Pandemic in (a) Madrid, (b) New York City (NYC), and (c) Mexico City. Actual data points, as officially reported, are shown using circles. Model predictions of new COVID-19 cases (yellow line) and simulation predictions of the cumulative number of symptomatic patients (blue line). The values of α and σ used in the simulations for each city are indicated within the corresponding pannels.

We also fitted the model to the pandemic progression observed in NYC. NYC is a densely populated area with a population mark of 8,400,000 inhabitants (i.e., nearly a Type-II city). Therefore, only a combination of social distancing and intensified testing can stop progression effectively (Table 3). Our results suggest that NYC has applied a strategy mostly based on aggressive testing.

Indeed, a scenario based on a sustained level of social distancing of 27% efficacy (σ=0.27; effective reduction of 30% in demographic density) and a massive testing effort (α=0.98) is the combination that better recapitulates the actual pandemic evolution in NYC. NYC is, therefore, a remarkable example of the efficacy of massive diagnosis and quarantine, as a city that was able to stop COVID-19 progression at a cumulative count of ∼250,000 symptomatic subjects, which is about 25% of the maximum expected. In perspective, with an effective distancing of 27% and no aggressive testing (i.e., α=0.50 instead of α=0.98), the peak of bed occupancy in the city would have been at least three-fold higher, causing a total collapse of the hospital system. Our results also show that demographic density is a key factor in COVID-19 progression. Note that the testing effort in NYC was more intense than in Madrid. However, NYC has double the demographic density of Madrid, resulting in a higher number of those infected than in Madrid (250,000 versus 70,000).

While Madrid and NYC have transitioned through the pandemic peak and successfully extinguished the pandemic, Mexico City is still struggling to contain the pandemics. Several mathematical models have been used to forecast pandemic scenarios for Mexico City [40–42]. However, accurately predicting the progression of COVID-19 and the occurrence of the pandemic peak in the capital of Mexico has been challenging. Mexico City has a demographic density of 1.69 times that of Madrid and a population of more than 15,000 within the city limits. In terms of demographic density, Mexico City lies between a Type-I and Type-II city, possessing a demographic density 1.69 times greater than Madrid. However, in terms of population, Mexico City is double that of NYC. The Mexico City Metropolitan Area, which comprehends the city and its surrounding municipalities in Estado de México, houses 25,000,000 inhabitants. From the scenarios simulated before (Figure 4), we may readily infer that only a combination of aggressive social distancing and massive testing effort can stop pandemic progression in Mexico City. Mexican public health officials have publicly stated their decision to deemphasize diagnostics, instead privileging social distancing in the entire Mexican territory. Figure 4C compares the actual evolution of the number of cases in Mexico City versus the prediction of our model for a scenario where social distancing is set at σ=0.75 (75% effective reduction on demographic density) and the testing effort at α=0.10% (only 10% of infected individuals are effectively quarantined after being diagnosed). The Mexican strategy has been successful in delaying the pandemic’s progression, but not in bending the slope of the epidemic curve. As a result, Mexico City is steadily positioned in a pandemic plateau, with a nearly constant number of cases per day (600 cases), which is consistent with journalistic information and official data (https://coronavirus.gob.mx/datos/)[38].

## Concluding remarks

Here we introduce a mathematical model, based on demographic and clinical data that enables evaluating the relative benefit of social distancing and massive testing. Using this simple model, we investigate scenarios of COVID-19 evolution in two types of representative cities (i.e., 3,500,000 inhabitants and 5,000 hab km^2^, and 5.0 × 10^6^ inhabitants and 10,000 hab km^2^).

Our modeling simulations show that for Type-I cities, extreme and sustained social distancing (i.e., effectively decreasing the demographic density by 60% or more) may extinguish the pandemic progression. However, in Type-II cities (or larger), only combinations of social distancing and aggressive testing can effectively control the pandemic evolution. This finding has relevant implications for the planning of countermeasures for the effective containment of pandemic progression and for the design/redesign of urban areas. The concept of sustainability and cost-effectiveness of densely populated urban areas has to be revisited. Our results make explicit that large cities are highly vulnerable to epidemic crisis.

In principle, this model can be adapted to any urban area by setting the population and the demographic density. The predictions on the evolution of COVID-19 based on this mathematical model could represent important tools for designing and/or evaluating countermeasures.

## Data Availability

All relevant data and programs to reproduce the information presented here is available at the manuscript or as supplementary material.

## Acknowledgments

MMA and GTdS acknowledge the funding received from CONACyT (Consejo Nacional de Ciencia y Tecnología, México) and Tecnológico de Monterrey.

## Author contributions

MMA and GTdS collected and analyzed epidemiology data. MMA formulated the model and ran the simulations. MMA and GTdS wrote the manuscript. Both authors reviewed and approved the manuscript.

## Supporting Information

**S1 File**. Excel file that contains mathematical modelling formulation in Excel

